# Lymphocyte subset alterations with disease severity, imaging manifestation, and delayed hospitalization in COVID-19 patients

**DOI:** 10.1101/2020.08.25.20181321

**Authors:** Daxian Wu, Xiaoping Wu, Jiansheng Huang, Qunfang Rao, Qi Zhang, Wenfeng Zhang

**Author notes:** **Correspondence** Wenfeng Zhang, The First Affiliated Hospital, Nanchang University, No.17 Yongwai Street, Donghu District, 330006 Nanchang, Jiangxi Province, China.

## Abstract

**Background:** COVID-19 continuously threated public health heavily. Present study aimed to investigate the lymphocyte subset alterations with disease severity, imaging manifestation, and delayed hospitalization in COVID-19 patients.

**Methods:** Lymphocyte subsets was classified using flow cytometry with peripheral blood collected from 106 patients.

**Results:** Multivariate logistic regression showed that chest tightness, lymphocyte count, and γ-glutamyl transpeptidase were the independent predictors for severe COVID-19. The T cell, CD4^+^ T cell and B cell counts in severe patients were significantly lower than that in mild patients (p = 0.004, 0.003 and 0.046, respectively). Only the T cell count was gradually decreased with the increase of infiltrated quadrants of lesions in computed tomography (CT) (p = 0.043). The T cell, CD4^+^ T cell, and CD8^+^ T cell counts were gradually decreased with the increase of infiltrated area of the maximum lesion in CT (p = 0.002, 0.003, 0.028; respectively). The T cell count, as well as CD4^+^ T cell, CD8^+^ T cell, and NK cell counts were gradually decreased with the increased delayed hospitalization (p = 0.003, 0.002, 0.013, and 0.012; respectively). The proportion of T cell was gradually decreased but B cell was gradually increased with the increased delayed hospitalization (p = 0.031 and 0.003, respectively).

**Conclusion:** T cell and CD4^+^ T cell counts negatively correlated with disease severity, CT manifestation and delayed hospitalization. CD4^+^ T cell was mainstay of immunity response in COVID-19 patients.

## INTRODUCTION

Coronavirus disease 2019 (COVID-19) is a newly emerged viral pneumonia caused by the severe acute respiratory syndrome coronavirus 2 (SARS-CoV-2) ^[1]^. COVID-19 is highly contagious and has become pandemic quickly. Innate and adaptive immune responses were activated in COVID-19 patients. However, uncontrolled innate and adaptive immune responses may lead to locally and systemically tissue damage, which leads to severe COVID-19 with more frequent intubation and mechanical ventilation, and high mortality. Exhausted lymphocytes were a feature of severe COVID-19 ^[2-4]^. Recently, the alterations of lymphocyte subsets in COVID-19 patients had attracted the attention of researchers. An overall decline of lymphocyte subsets including CD4^+^ T cells, CD8^+^ T cells, B cells, and NK cells has been reported in severe and deceased COVID-19 patients ^[5, 6]^. However, vary patterns of lymphocyte subsets abnormality in severe COVID-19 patients also have been demonstrated by other studies ^[7-10]^. Reports involving the change of CD4^+^ to CD8^+^ T cells ratio were also inconsistent ^[9, 11, 12]^. Thus, the reported patterns of lymphocyte subsets in patients with COVID-19 were diverse and controversial, and necessitated to clear more. Additionally, the correlations of lymphocyte subsets with the lesion manifestation in computed tomography (CT) and with the time from onset to hospitalization (TOH) of patients were also not be well documented.

Here, we first investigated alterations of lymphocyte subsets in severe COVID-19 patients. Then, we observed the correlations of lymphocyte subsets with the number, quadrant, and area of lesions in CT. Finally, we investigated the impacts of TOH of patients on lymphocyte subsets.

## METHOD

### Patients

One hundred and six COVID-19 patients who were confirmed by positive RNA of SARS-CoV-2 using throat swab specimens were recruited from June 23, 2020 to February 29, 2020 at the First Affiliated Hospital, Nanchang University. Patients were stratified at their admission, severe COVID-19 was diagnosed according the guideline of the American Thoracic Society and Infectious Diseases Society of America ^[13]^. Cases not meeting the criteria were classified as mild COVID-19. All procedures followed were in accordance with the Ethics Committees of the First Affiliated Hospital, Nanchang University, and with the Helsinki Declaration of 1975, as revised in 2000. Informed consent was obtained from all patients for being included in the study.

### Data acquisition

Data on the demography, epidemiology, symptoms and signs, laboratory tests, as well as radiography findings were extracted from electronic medical records using a predesigned datasheet. All laboratory tests were conducted in the Central Clinical Laboratory of the First Affiliated Hospital, Nanchang University and were adopted if they were performed with fasting blood samples at patients’ admission.

### Flow cytometry

Anticoagulated peripheral blood samples with EDTA were collected from COVID-19 patients at their admission and tested within 6 hours. Lymphocyte subsets was performed by BD FACSCanto™ II Flow Cytometer (BD Biosciences, Shanghai, China). Anti-CD3 was conjugated by fluorescein isothiocyanate (FITC), anti-CD4, anti-CD8, anti-CD19 were conjugated by allophycocyanin (APC), phycoerythrin (PE), and APC respectively, anti-CD16 and anti-CD56 conjugated by PE (340499 and 340500, BD Biosciences). All tests were performed according to the manufacturer’s instructions.

### Statistics

Statistical analysis was performed with SPSS 25.0 (SPSS, Inc., Chicago, USA) and MedCalc (MedCalc Software Ltd, Ostend, Belgium). Continuous data were expressed as the mean ± standard deviations or medians with quartile (P25-P75) and categorical data were expressed as numbers (%). Student’s t-test or the Mann–Whitney U test as appropriate were used for continuous data, and the χ2 or Fisher’s tests were used for categorical data. Rank correlation was analysed using the Spearman method. A p values of less than 0.05 was considered statistically significant. Independent risk factors were identified using multivariate logistic regression according to the forward Wald method, with entry and removal probabilities of 0.05 and 0.10, respectively.

## RESULTS

### Baseline clinical characteristics of patients with COVID-19

The age of patients with COVID-19 was 46.17 ± 14.39, and 60.4% patients were male. The mean of TOH was 5 days. Among the 106 COVID-19 patients, 83 (78.3%) patients had a clear exposure history, and 33 (31.1%) patients had one or more comorbidities. The most frequent comorbidity was bacterial infection (11.3%), followed by diabetes (9.4%) and hypertension (7.5%). As expected, fever (91.5%), dry cough (43.4%), and chest tightness (32.1%) were the top three frequent symptoms. Chills (20.8%), fatigue (19.8%), and sore throat (18.9%) were also common, but rhinorrhea or rhinobyon (5.7%), diarrhea (6.6%), and myalgia (7.5%) were relatively rare in COVID-19 patients (Table 1).

**Table 1.**
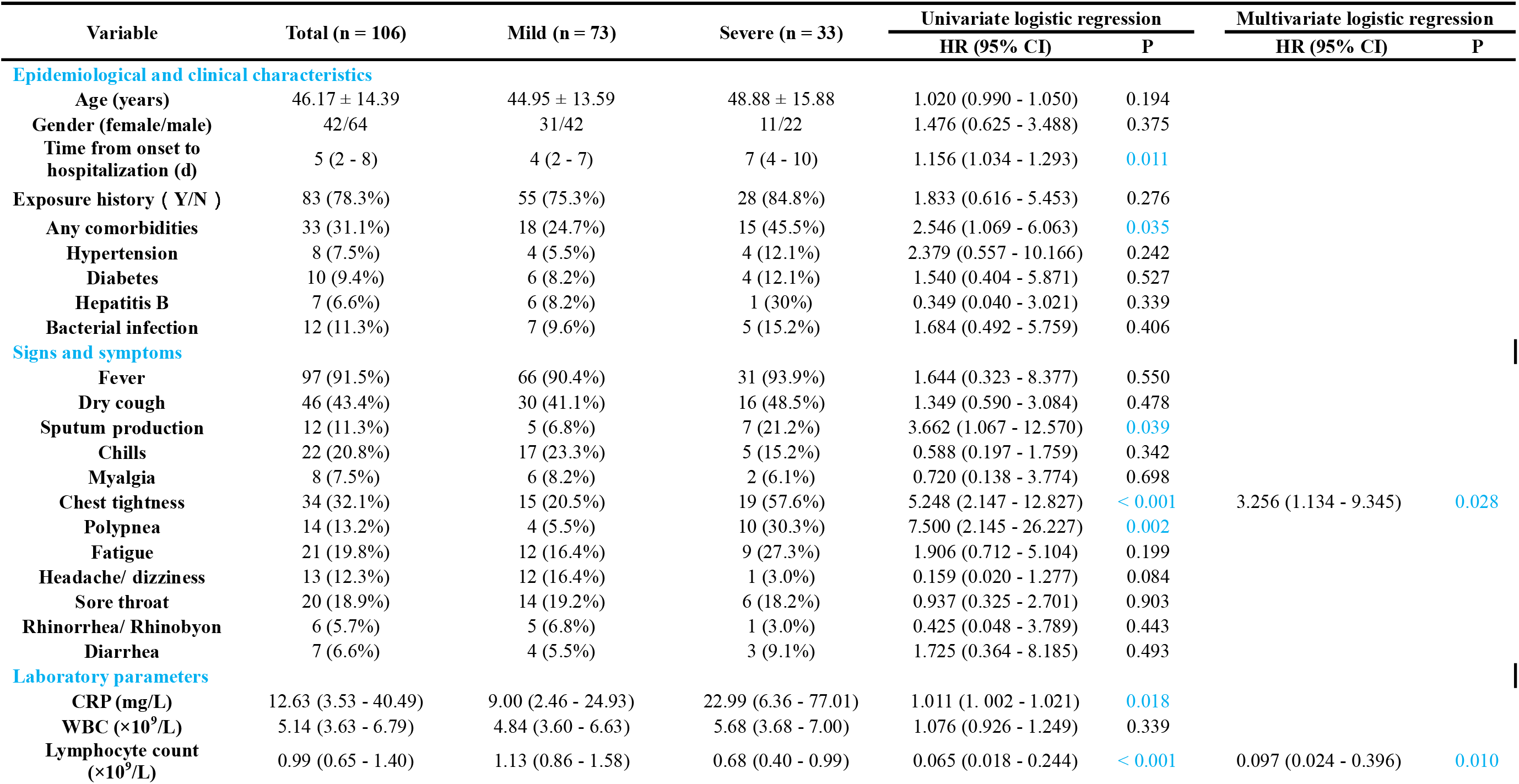
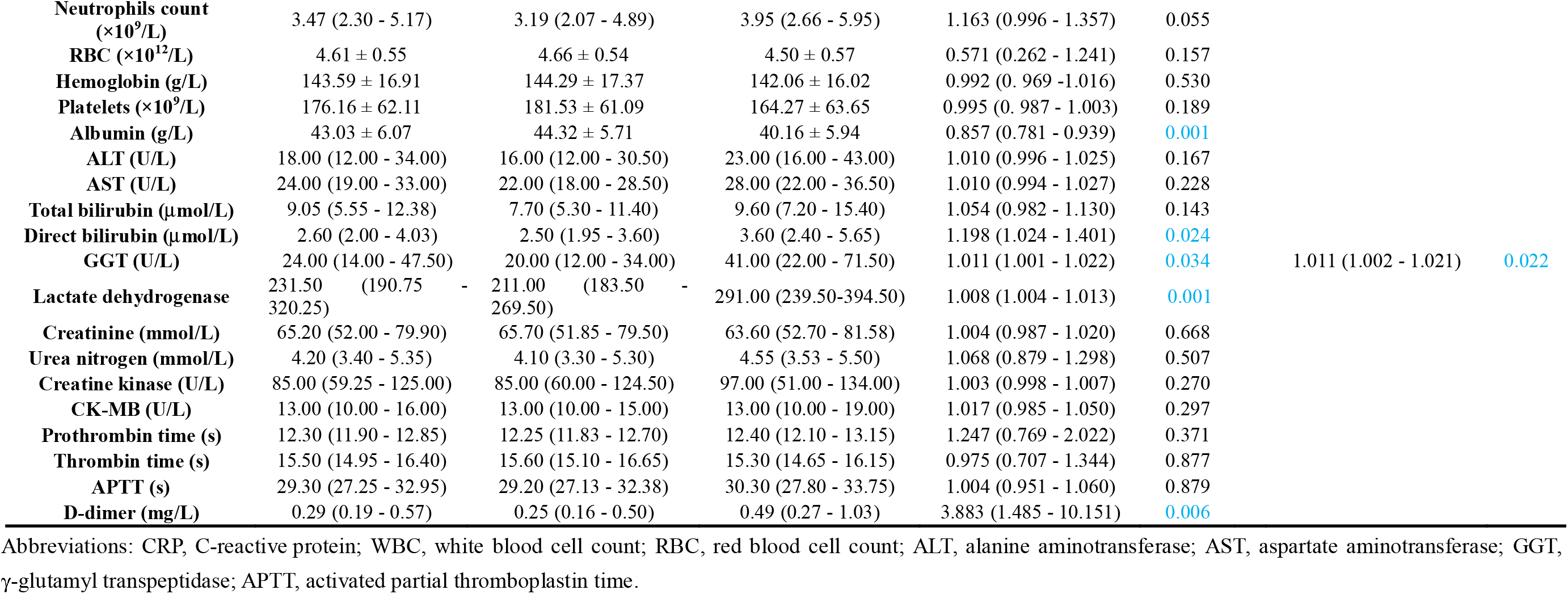
Characteristics at admission of the patients with COVID-19

| Variable | Total (n = 106) | Mild (n = 73) | Severe (n = 33) | Univariate logistic regression |  | Multivariate logistic regression |  |
| --- | --- | --- | --- | --- | --- | --- | --- |
|  |  |  |  | HR (95% CI) | P | HR (95% CI) | P |
| Epidemiological and clinical characteristics |  |  |  |  |  |  |  |
| Age (years) | 46.17 ± 14.39 | 44.95 ± 13.59 | 48.88 ± 15.88 | 1.020 (0.990 - 1.050) | 0.194 |  |  |
| Gender (female/male) | 42/64 | 31/42 | 11/22 | 1.476 (0.625 - 3.488) | 0.375 |  |  |
| Time from onset to hospitalization (d) | 5 (2 - 8) | 4 (2 - 7) | 7 (4 - 10) | 1.156 (1.034 - 1.293) | 0.011 |  |  |
| Exposure history ( Y/N ) | 83 (78.3%) | 55 (75.3%) | 28 (84.8%) | 1.833 (0.616 - 5.453) | 0.276 |  |  |
| Any comorbidities | 33 (31.1%) | 18 (24.7%) | 15 (45.5%) | 2.546 (1.069 - 6.063) | 0.035 |  |  |
| Hypertension | 8 (7.5%) | 4 (5.5%) | 4 (12.1%) | 2.379 (0.557 - 10.166) | 0.242 |  |  |
| Diabetes | 10 (9.4%) | 6 (8.2%) | 4 (12.1%) | 1.540 (0.404 - 5.871) | 0.527 |  |  |
| Hepatitis B | 7 (6.6%) | 6 (8.2%) | 1 (30%) | 0.349 (0.040 - 3.021) | 0.339 |  |  |
| Bacterial infection | 12 (11.3%) | 7 (9.6%) | 5 (15.2%) | 1.684 (0.492 - 5.759) | 0.406 |  |  |
| Signs and symptoms |  |  |  |  |  |  |  |
| Fever | 97 (91.5%) | 66 (90.4%) | 31 (93.9%) | 1.644 (0.323 - 8.377) | 0.550 |  |  |
| Dry cough | 46 (43.4%) | 30 (41.1%) | 16 (48.5%) | 1.349 (0.590 - 3.084) | 0.478 |  |  |
| Sputum production | 12 (11.3%) | 5 (6.8%) | 7 (21.2%) | 3.662 (1.067 - 12.570) | 0.039 |  |  |
| Chills | 22 (20.8%) | 17 (23.3%) | 5 (15.2%) | 0.588 (0.197 - 1.759) | 0.342 |  |  |
| Myalgia | 8 (7.5%) | 6 (8.2%) | 2 (6.1%) | 0.720 (0.138 - 3.774) | 0.698 |  |  |
| Chest tightness | 34 (32.1%) | 15 (20.5%) | 19 (57.6%) | 5.248 (2.147 - 12.827) | < 0.001 | 3.256 (1.134 - 9.345) | 0.028 |
| Polypnea | 14 (13.2%) | 4 (5.5%) | 10 (30.3%) | 7.500 (2.145 - 26.227) | 0.002 |  |  |
| Fatigue | 21 (19.8%) | 12 (16.4%) | 9 (27.3%) | 1.906 (0.712 - 5.104) | 0.199 |  |  |
| Headache/ dizziness | 13 (12.3%) | 12 (16.4%) | 1 (3.0%) | 0.159 (0.020 - 1.277) | 0.084 |  |  |
| Sore throat | 20 (18.9%) | 14 (19.2%) | 6 (18.2%) | 0.937 (0.325 - 2.701) | 0.903 |  |  |
| Rhinorrhea/ Rhinobyon | 6 (5.7%) | 5 (6.8%) | 1 (3.0%) | 0.425 (0.048 - 3.789) | 0.443 |  |  |
| Diarrhea | 7 (6.6%) | 4 (5.5%) | 3 (9.1%) | 1.725 (0.364 - 8.185) | 0.493 |  |  |
| Laboratory parameters |  |  |  |  |  |  |  |
| CRP (mg/L) | 12.63 (3.53 - 40.49) | 9.00 (2.46 - 24.93) | 22.99 (6.36 - 77.01) | 1.011 (1.002 - 1.021) | 0.018 |  |  |
| WBC (×10<sup>9</sup>/L) | 5.14 (3.63 - 6.79) | 4.84 (3.60 - 6.63) | 5.68 (3.68 - 7.00) | 1.076 (0.926 - 1.249) | 0.339 |  |  |
| Lymphocyte count (×10<sup>9</sup>/L) | 0.99 (0.65 - 1.40) | 1.13 (0.86 - 1.58) | 0.68 (0.40 - 0.99) | 0.065 (0.018 - 0.244) | < 0.001 | 0.097 (0.024 - 0.396) | 0.010 |
| **Neutrophils count**<br>($\times 10^9/L$) | 3.47 (2.30 - 5.17) | 3.19 (2.07 - 4.89) | 3.95 (2.66 - 5.95) | 1.163 (0.996 - 1.357) | 0.055 | | |
| **RBC** ($\times 10^{12}/L$) | 4.61 ± 0.55 | 4.66 ± 0.54 | 4.50 ± 0.57 | 0.571 (0.262 - 1.241) | 0.157 | | |
| **Hemoglobin** (g/L) | 143.59 ± 16.91 | 144.29 ± 17.37 | 142.06 ± 16.02 | 0.992 (0.969 - 1.016) | 0.530 | | |
| **Platelets** ($\times 10^9/L$) | 176.16 ± 62.11 | 181.53 ± 61.09 | 164.27 ± 63.65 | 0.995 (0.987 - 1.003) | 0.189 | | |
| **Albumin** (g/L) | 43.03 ± 6.07 | 44.32 ± 5.71 | 40.16 ± 5.94 | 0.857 (0.781 - 0.939) | 0.001 | | |
| **ALT** (U/L) | 18.00 (12.00 - 34.00) | 16.00 (12.00 - 30.50) | 23.00 (16.00 - 43.00) | 1.010 (0.996 - 1.025) | 0.167 |  |  |
| **AST** (U/L) | 24.00 (19.00 - 33.00) | 22.00 (18.00 - 28.50) | 28.00 (22.00 - 36.50) | 1.010 (0.994 - 1.027) | 0.228 |  |  |
| **Total bilirubin** ($\mu\text{mol/L}$) | 9.05 (5.55 - 12.38) | 7.70 (5.30 - 11.40) | 9.60 (7.20 - 15.40) | 1.054 (0.982 - 1.130) | 0.143 | | |
| **Direct bilirubin** ($\mu\text{mol/L}$) | 2.60 (2.00 - 4.03) | 2.50 (1.95 - 3.60) | 3.60 (2.40 - 5.65) | 1.198 (1.024 - 1.401) | 0.024 | | |
| **GGT** (U/L) | 24.00 (14.00 - 47.50) | 20.00 (12.00 - 34.00) | 41.00 (22.00 - 71.50) | 1.011 (1.001 - 1.022) | 0.034 | 1.011 (1.002 - 1.021) | 0.022 |
| **Lactate dehydrogenase** | 231.50 (190.75 - 320.25) | 211.00 (183.50 - 269.50) | 291.00 (239.50-394.50) | 1.008 (1.004 - 1.013) | 0.001 |  |  |
| **Creatinine** (mmol/L) | 65.20 (52.00 - 79.90) | 65.70 (51.85 - 79.50) | 63.60 (52.70 - 81.58) | 1.004 (0.987 - 1.020) | 0.668 |  |  |
| **Urea nitrogen** (mmol/L) | 4.20 (3.40 - 5.35) | 4.10 (3.30 - 5.30) | 4.55 (3.53 - 5.50) | 1.068 (0.879 - 1.298) | 0.507 |  |  |
| **Creatine kinase** (U/L) | 85.00 (59.25 - 125.00) | 85.00 (60.00 - 124.50) | 97.00 (51.00 - 134.00) | 1.003 (0.998 - 1.007) | 0.270 |  |  |
| **CK-MB** (U/L) | 13.00 (10.00 - 16.00) | 13.00 (10.00 - 15.00) | 13.00 (10.00 - 19.00) | 1.017 (0.985 - 1.050) | 0.297 |  |  |
| **Prothrombin time** (s) | 12.30 (11.90 - 12.85) | 12.25 (11.83 - 12.70) | 12.40 (12.10 - 13.15) | 1.247 (0.769 - 2.022) | 0.371 |  |  |
| **Thrombin time** (s) | 15.50 (14.95 - 16.40) | 15.60 (15.10 - 16.65) | 15.30 (14.65 - 16.15) | 0.975 (0.707 - 1.344) | 0.877 |  |  |
| **APTT** (s) | 29.30 (27.25 - 32.95) | 29.20 (27.13 - 32.38) | 30.30 (27.80 - 33.75) | 1.004 (0.951 - 1.060) | 0.879 |  |  |
| **D-dimer** (mg/L) | 0.29 (0.19 - 0.57) | 0.25 (0.16 - 0.50) | 0.49 (0.27 - 1.03) | 3.883 (1.485 - 10.151) | 0.006 |  |  |

Abbreviations: CRP, C-reactive protein; WBC, white blood cell count; RBC, red blood cell count; ALT, alanine aminotransferase; AST, aspartate aminotransferase; GGT, γ-glutamyl transpeptidase; APTT, activated partial thromboplastin time.

### Independent indictors for severe COVID-19

As shown in Table 1, univariate logistic regression indicated that the TOH of severe patients was significantly longer than of mild patients [7 (4 - 10) vs. 4 (2 - 7) days; p = 0.011]. The frequency of comorbidities in severe patients was higher than mild patients (45.5% vs. 24.7%; p = 0.035). The frequency of sputum production, chest tightness, and polypnea was more higher in severe patients than mild patients (all p < 0.05). The level of C-reactive protein, as well as levels of direct bilirubin, γ-glutamyl transpeptidase, lactate dehydrogenase, and D-dimer were significantly higher in severe patients than mild patients (all p < 0.05). However, the levels of lymphocyte count and albumin were significantly lower in severe cases compared to mild cases. Multivariate logistic regression showed that chest tightness, lymphocyte count, and γ-glutamyl transpeptidase were the independent indictors to predict severe COVID-19.

### Lymphocyte subsets in severe COVID-19 patients

Giving the lymphocyte count was an important indicator to predict severe COVID-19, we further investigated the alteration of lymphocyte subsets in patients with severe COVID-19. As shown in Figure 1A, the T cell count in severe patients was significantly lower than that in mild patients [487.00 (291.50, 819.50) vs. 766.00 (525.50, 1036.50) /μL; p = 0.004]. The CD4^+^ T cell and B cell counts in severe patients were also significantly lower than that in mild patients [272.00 (177.00, 497.50) vs. 455.00 (283.50, 612.50) and 92.00 (57.50, 160.00) vs. 136.00 (82.50, 213.00) /μL; p = 0.003 and 0.046, respectively]. There is no significant difference for CD8^+^ T cell or NK cell count between severe and mild patients. The difference of CD4^+^ To CD8+ ratio between severe and mild patients were not significant. No significant difference for proportion of lymphocyte subset was observed between severe and mild patients (Figure 1B).

**Fig. 1.**
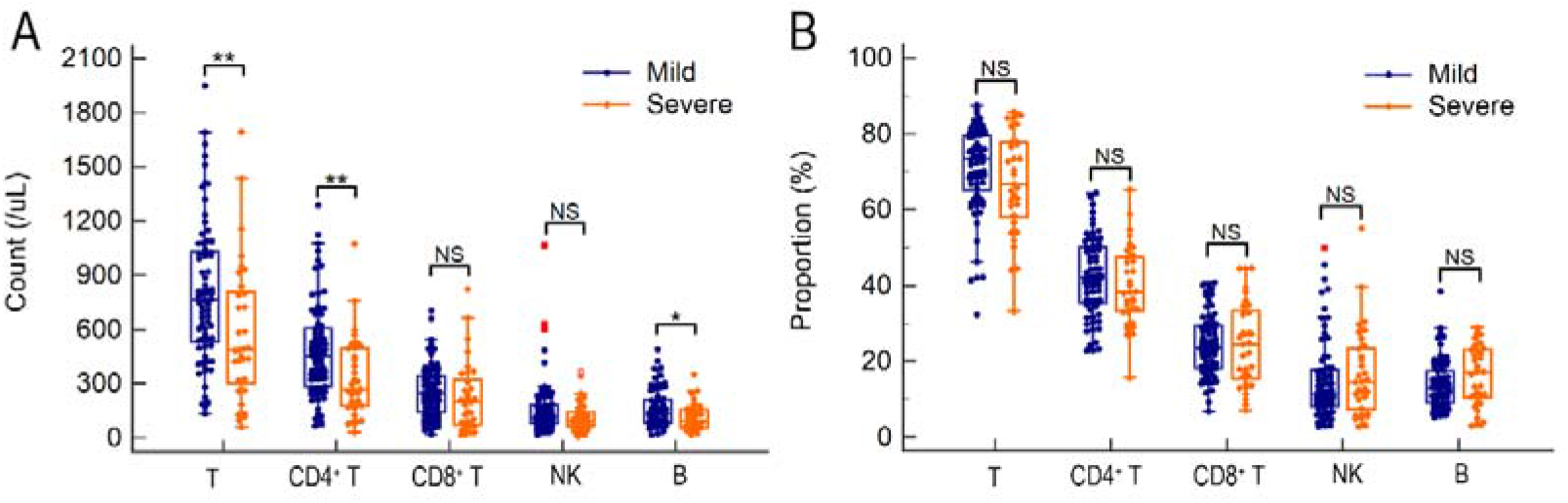
The lymphocyte subset alterations in severe COVID-19 patients

### Lymphocyte subsets alterations with CT manifestation

In order to assess the manifestation of lesions in CT, present study simply scored the number, quadrant, and area of lesions. For the number of lesions, patients were classified to 3 subgroups named patients with no lesion, ≤3 lesions, and >3 lesions. For the quadrant of lesions, patients were classified to 3 subgroups, that is, patients with no quadrant, ≤3 quadrants, and >3 quadrants. For the area of the maximum lesion, patients were classified no infiltration when there is no lesion in CT. patients with minor and major infiltration were classified when the area of the maximum lesion were ≤100 cm^2^ and >100 cm^2^ respectively. As shown in Figure 2A-C, the lymphocyte counts were gradually decreased with the increased number, quadrant, or area of lesions (p = 0.002, 0.002, and <0.001 respectively). No significant trend of absolute count of any lymphocyte subset was observed with the increase of lesion number (Figure 3 A). Only the T cell count was gradually decreased with the increase of infiltrated quadrants [919.50 (699.00, 1274.75), 715.00 (452.00, 1020.50), and 607.00 (398.00, 912.00) /μL, p = 0.043] (Figure 3B). The T cell, CD4^+^ T cell, and CD8^+^ T cell counts were gradually decreased with the increase of infiltrated area [919.50 (699.00, 1274.75), 724.00 (487.00, 1021.00), and 494.00 (263.00, 796.00)/μL, p = 0.002 for T cell; 547.00 (428.75, 835.00), 411.00 (266.00, 587.00), and 325.00 (173.00, 501.00) /μL, p = 0.003 for CD4^+^ T cell; 292.00 (217.00, 351.75), 228.00 (128.00, 359.00), and 158.00 (83.50, 283.00) /μL, p = 0.028 for CD8^+^ T cell; respectively] (Figure 3C. However, the trend of CD4^+^ To CD8^+^ ratio was not significant no matter with the increase of the number, quadrant, or the area of lesions. For proportion, there is also no significant trend of any lymphocyte subset with aggravated CT manifestation (Figure 3D-F).

**Fig. 2.**
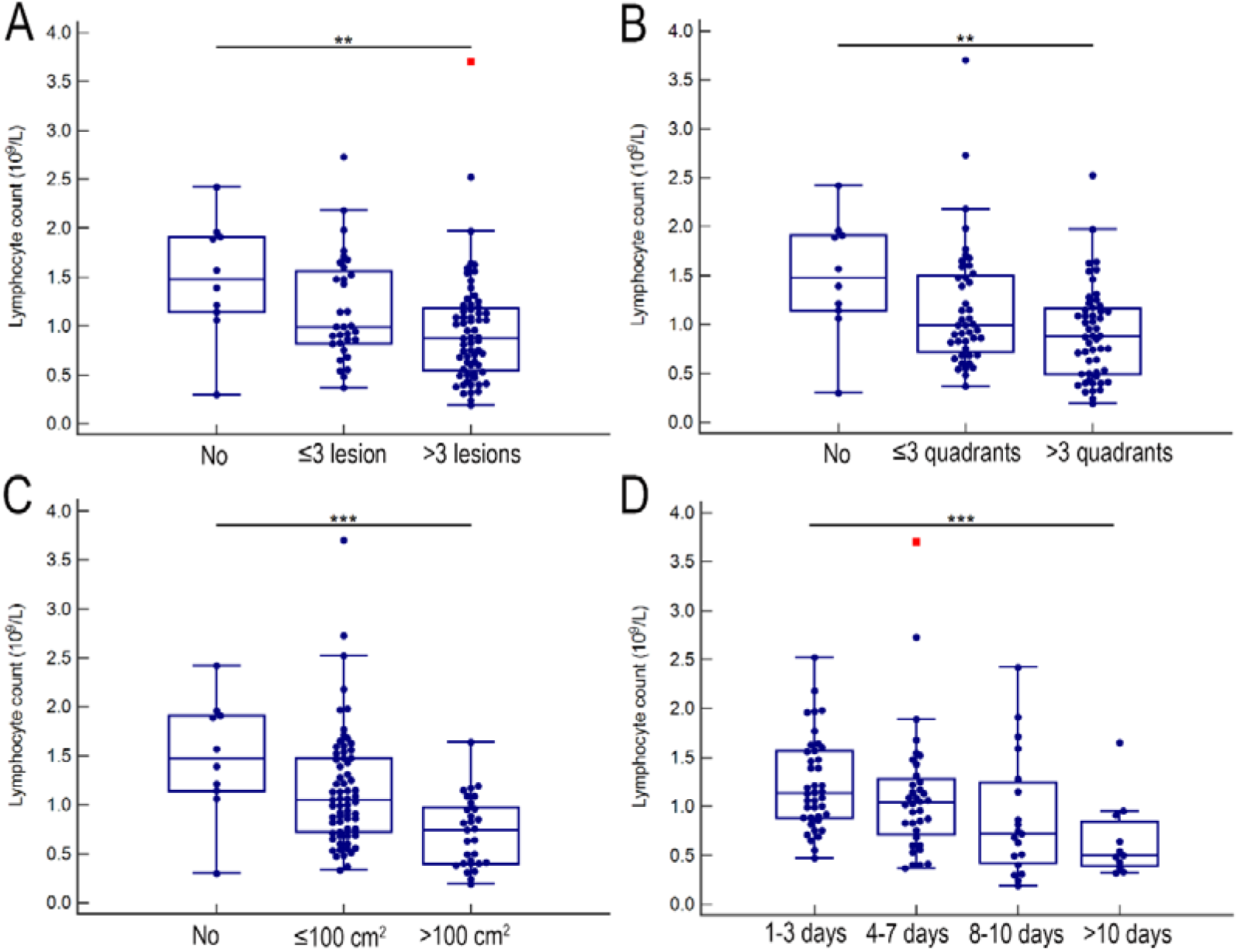
The total lymphocyte cell count with aggravated CT manifestation and increased delayed hospitalization

**Fig. 3.**
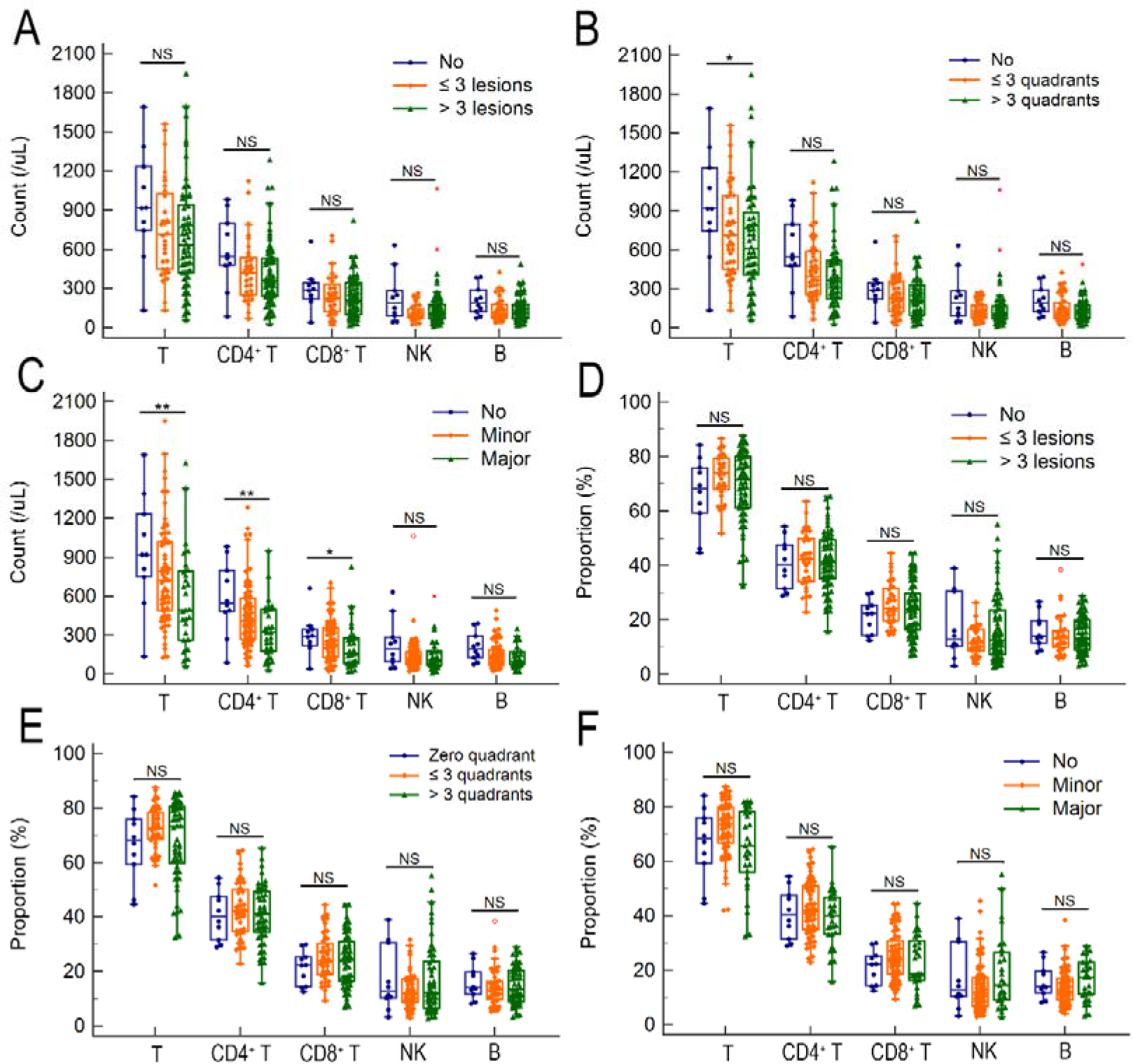
The lymphocyte subset alterations with aggravated CT manifestation

### Lymphocyte subsets alterations with TOH

The lymphocyte counts were gradually decreased with the increased TOH (p < 0.001) (Figure 2D). Lymphocyte subsets analysis showed that the T cell count, as well as CD4^+^ T cell, and CD8^+^ T cell counts were gradually decreased with the increased TOH [736.50 (541.50, 1022.00), 764.00 (495.00, 1016.50), 512.00 (227.00, 950.00), 425.00 (136.00, 694.00), p = 0.003 for T cell count; 481.00 (303.25, 596.00), 404.00 (243.25, 546.00), 259.00 (131.00, 707.00), 272.00 (96.00, 502.00), p = 0.002 for CD4^+^ T cell count; 250.00 (140.75, 345.75), 238.00 (158.75, 378.00), 200.00 (63.00, 370.00), 75.00 (45.00, 180.00), p = 0.013 for CD8^+^ T cell count]. The NK cell count was also gradually decreased with the increased TOH [120.50 (88.00, 218.75), 111.50 (86.25, 175.25), 93.00 (48.00, 180.00), 69.00 (61.00, 110.00), p = 0.012] (Figure 4A). There is no significant trend of CD4^+^ To CD8^+^ ratio with the delayed hospitalization. The proportion of T cell was gradually decreased with the increased TOH [74.25 (67.33, 79.35), 75.85 (62.93, 80.88), 67.20 (56.60, 79.30), 61.70 (54.00, 68.50), p = 0.031], but the proportion of B cell was gradually increased with the increased TOH [11.70 (8.85, 17.05), 11.25 (7.48, 17.35), 17.80 (13.60, 23.50), 17.40 (13.40, 26.30), p = 0.003] (Figure 4B).

**Fig. 4.**
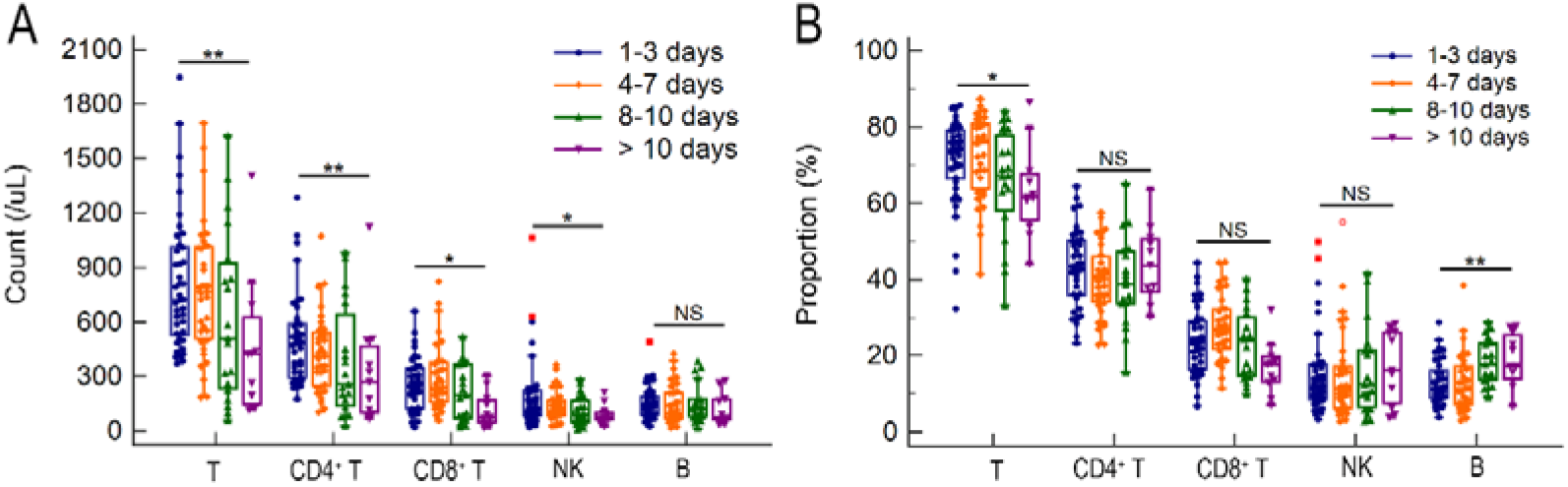
The lymphocyte subset alterations with increased delayed hospitalization

## DISCUSSION

COVID-19 continuously threated public health heavily, which required more bench and clinic studies to more profoundly profile this disease. The common symptoms in COVID-19 patients, in accordance with previous reports ^[2, 14, 15]^, were fever, and followed by dry cough, and chest tightness. Similar with studies investigated in other area ^[16, 17]^, the age of patients in this study was dramatically younger than those in Wuhan. Notably, the frequencies of hypertension and diabetes for COVID-19 patients in this study were also much lower than patients in Wuhan^[2, 14, 18]^. The underlying reason may interpret by the lower age. In addition, present study showed that there are 11.3% patients accompanying with bacterial infection, which suggested more attention should be paid to the evaluation of bacterial infection on patients’ admission.

Lymphocytes and their subsets play a key role in the maintenance of immune function. Complex immune dysregulation has been found in COVID-19 patients ^[19]^. Currently, the change patterns of lymphocyte subsets were not conclusive. It is reported ^[5, 6]^ that an whole decline of lymphocyte subsets including CD4^+^ and CD8^+^ T cells, B cells, and NK cells were presented in severe and deceased COVID-19 patients. Liu et al. ^[10]^ suggested CD8^+^ T cell count was significant decreased in severe COVID-19 patients than mild patients at the time point of disease onset and 7-9 days later, but their difference in CD4^+^ T cell count was not significant at any time point. studies ^[7-9]^ also indicated decreased CD4+ and CD8^+^ T cells were correlated with disease severity of COVID-19, but there is no difference for the level of B or NK cell between severe and mild COVID-19 patients. Additionally, reports involving the change of CD4^+^ to CD8^+^ T cells ratio were also inconsistent ^[9, 11, 12]^. In this study, we found the significant percentage change of lymphocyte subsets is rare no matter in patients with severer clinical type, more extensive CT manifestation, or with longer delayed hospitalization. These findings were agreement with previous studies ^[4, 7]^. It is noteworthy that the T cell and CD4^+^ T cell but not CD8^+^ T cell were significantly decreased in severe COVID-19 patents, which suggested that CD4^+^ T cell but not CD8^+^ T cell play more important role in immunity response to SARS-CoV-2 infection. Studies using SARS-CoV or MERS-CoV infected mouse demonstrated that depletion of CD4^+^ T cells but not CD8^+^ T cells would lead to delayed clearance of virus and enhanced immune-mediated pneumonitis ^[20, 21]^. Similarly, high-level CD4+ but not the CD8^+^ T cell response was also observed in SARS patients ^[22]^. What is more, the significantly decreased B cell in severe COVID-19 patents indicated that humoral immunity has been attenuated in antiviral response of SARS-CoV-2. It has reported ^[23-25]^ that T-helper type 1 (Th1), T-helper type 2 (Th2), and regulatory T cells were varying degrees of activated in peripheral blood from critical COVID-19 patients after stimulation with specific antigen of SARS-CoV-2. It can be speculated all the CD4^+^ T cell subgroups were exhaust in blood of critical COVID-19 patients for the severely damaged lymphoid organs and/or exudation of circulating lymphocytes into lung ^[9]^, although the alteration of CD4^+^ T cell subsets warrants further investigation.

With regard to lymphocyte subset changes with CT manifestation, present study found that the total lymphocyte counts were gradually decreased with the increased number, infiltrated quadrants of lesions, and the area of the maximum lesion. T cell counts were gradually decreased with the increase of infiltrated quadrants of lesions and the area of the maximum lesion rather than the increased lesion number. Gradually decreased CD4+ and CD8^+^ T cell counts were only observed with increased area of the maximum lesion. Those results revealed that the area of the maximum lesion was closer correlated with the count of lymphocyte subsets and was more appropriate to estimate the severity of COVID-19.

The alteration of lymphocyte subsets with the delayed hospitalization has not been reported before, present study firstly observed their correlation and found that the total lymphocyte, T cell, CD4+ and CD8^+^ T cell counts were gradually decreased with the the increased TOH. This result indicated that the lymphatic organs will continue to be damaged as long as there is no intervention. Liver was a predominantly vulnerable extrapulmonary organ in patients with COVID-19, hepatic dysfunction was seen in 14-53% of cases and particularly in those with severe condition ^[26]^. Similar with the trends of lymphocyte subsets, present study also found the level of ALT was gradually elevated with the TOH (Figure 5), suggested that delayed hospitalization may cause more liver injure. Therefore, early hospitalization could avoid disease aggravation, more organ damage and the unnecessary use of scarce medical resources.

**Fig. 5.**
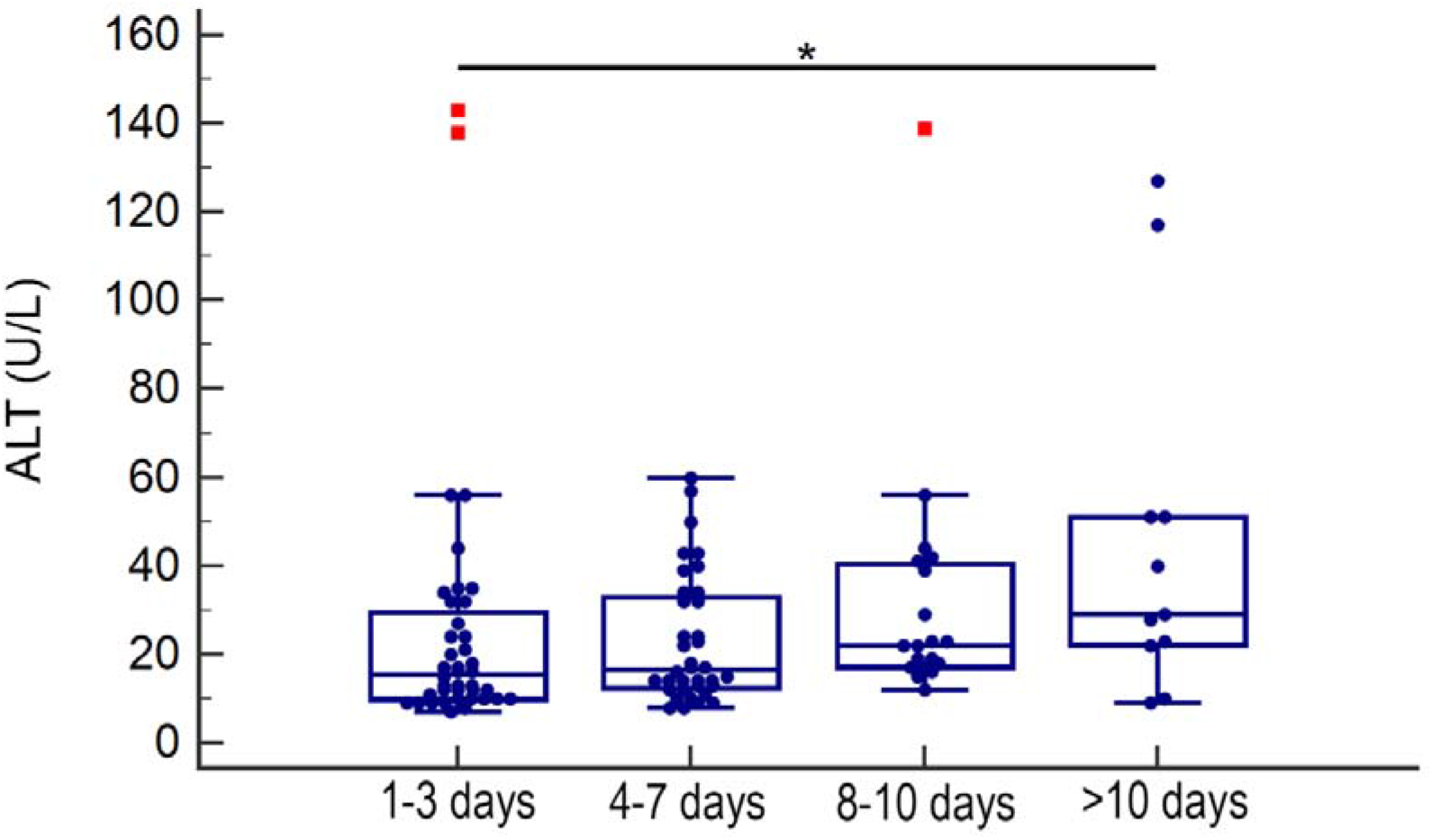
The alanine aminotransferase alterations with increased delayed hospitalization

There were several limitations in this study. First, the alteration of CD4^+^ T cell subsets was not investigated, although CD4^+^ T cell was demonstrated to be mainstay of immunity response to SARS-CoV-2 infection. Second, only 3 cases with TOH more than 14 days (TOH was 15, 15, and 16 days, respectively), the lymphocyte subset alterations of convalescence of COVID-19 patients was not seen in this study. More studies including patients with TOH more than 14 days need to investigate to observe lymphocyte subset alterations in whole natural history of the disease.

In conclusion, present study revealed independent predictors for severe COVID-19 and found CD4^+^ T cell was mainstay of immunity response to SARS-CoV-2 infection. Total lymphocyte, T cell and CD4^+^ T cell counts negatively correlated with disease severity, CT manifestation and delayed hospitalization. We believe these findings would provide some new insights in management of COVID-19.

## Data Availability

Please contact us via

## Declarations

### Ethics approval and consent to participate

All procedures followed were in accordance with the Ethics Committees of the First Affiliated Hospital, Nanchang University. Informed consent was obtained from all patients included in the study.

### Consent for publication

Not applicable

### Competing interests

The authors declare that they have no competing interests

### Funding

This work was funded by the National Science and Technology Major Project of China (2018ZX10302205), the Personnel Plan of Jiangxi Science and Technology Department (2016BCD40015), and the Project of Education Department of Jiangxi Province (GJJ170043).

### Authors’ contributions

Wenfeng Zhang were the guarantor of the submission. Daxian Wu, Xiaoping Wu, and Wenfeng Zhang designed the study. Jiansheng Huang, Qunfang Rao, and Qi Zhang enrolled the patients and collected the data. Daxian Wu and Wenfeng Zhang performed the statistical analysis of this work. Daxian Wu and Wenfeng Zhang analyzed and interpreted the data. Daxian Wu drafted the manuscript and Wenfeng Zhang provided critical revision of the manuscript. All authors approved the final version of the manuscript.

## Acknowledgements

Not applicable

## Availability of data and material

The datasets generated during and/or analysed during the current study are available from the corresponding author on reasonable request.

## Notes

### Competing Interest Statement

The authors have declared no competing interest.

### Author Declarations

The Ethics Committees of the First Affiliated Hospital, Nanchang University

